# Angiographic profile among patients with acute coronary syndrome in tertiary care centre: A retrospective cross-sectional study

**DOI:** 10.1101/2023.10.16.23297079

**Authors:** Shyam Raj Regmi, Ram Narayan Kurmi, Asraf Hussain, Dhiranjan Kumar Shah

## Abstract

**Background:** Acute coronary syndrome (ACS) is characterized by a variable presentation of vessel involvement, which significantly impacts morbidity and mortality rates. In this study, we aimed to investigate the angiographic findings in patients diagnosed with ACS.

**Materials and methods:** We conducted a retrospective cross-sectional study in the Cardiology Unit of the Department of Internal Medicine from January 1, 2022, to December 30, 2022, following ethical approval from the Institutional Review Committee (reference number: 079/080-161). We employed convenience sampling and calculated point estimates along with 95% confidence intervals.

**Results:** Among the 192 patients included in the study, 129 (67.18%) were male, with an average age of 61.29 ± 12.50 years. The most prevalent angiographic findings were triple vessel disease (TVD) in 77 (40.10%) cases and involvement of the left anterior descending artery (LAD) in 157 (81.77%) cases. ST-elevated myocardial infarction (STEMI) was present in 134 (89.23%) cases. Diabetes mellitus was the most common risk factor, present in 122 (63.54%) patients. Complications arose in 44 (22.91%) patients, with arrhythmia occurring in 18 (40.90%) cases. The mean hospital stay was 4.42 ± 1.87 days, and 10 (5.20%) patients unfortunately passed away during their hospitalization

**Conclusion:** Our study revealed that triple vessel disease and involvement of the left anterior descending artery were the most frequent angiographic findings in patients with ACS. Additionally, diabetes mellitus emerged as the predominant modifiable risk factor, potentially contributing to the prevalence of multiple vessel involvement.

## Introduction

Coronary artery disease (CAD) is a global health problem and is the leading cause of mortality and morbidity worldwide both in developed and developing countries.[1,2] Globally, South Asian countries have the highest incidence of coronary artery disease.[3] In Nepal Coronary Heart Disease related death reached 30559(18.7%) of total deaths with an Age-adjusted Death Rate of 158.35 per 100,000 of the population which ranks Nepal 41^st^ in the world according to WHO in 2017.[4]

In Nepal, the prevalence of CAD was 5.7%[5]. Percutaneous Coronary Intervention (PCI) was performed in limited hospital outside the capital city. Chitwan Medical College is the main referral tertiary care hospital in Central Terai. PCI was performed in patients with Acute Coronary Syndrome (ACS) confirmed by electrocardiographic (ECG) findings and cardiac biomarkers (cTnI).

The study aimed to determine the angiographic profile of patients with Acute Coronary Syndrome admitted to a tertiary care center.

## Materials and methods

This retrospective cross-sectional study was conducted in the Unit of Cardiology, Department of Internal Medicine, at Chitwan Medical College Teaching Hospital (CMCTH), from January 1, 2022 to December 30, 2022. The study was approved by the Institutional Review Committee (IRC) of CMCTH (Reference No-079/080-161). The data were reviewed from the Cardiology department from 15^th^ June 2023 to 22^nd^ June 2023 after ethical approval from the IRC. All patients above 18 years of age with coronary angiography admitted to the Coronary Care Unit (CCU) were included in the study. Patients without coronary angiography result or incomplete medical records were excluded from the study. Convenience sampling was used in this study. The sample size was calculated using following formula:

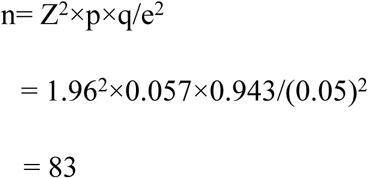

Where,

n= minimum require sample size

Z= 1.96 at 95% Confidence Interval (CI) p= prevalence of 5.7%[5]

q= 1-p

e= margin of error, 5%

The calculated sample size was 83. Taking 10% as the non-response rate, the required sample size was 91. However, total of 192 samples were taken for the study.

Acute STEMI was defined as chest pain of >20 min duration and ST-segment elevation >1 mm in at least two contiguous precordial standard limb leads or >2 mm in at least two contiguous precordial leads presenting within 7 days of symptoms with positive cardiac biomarkers.

Dyslipidemia was defined as the presence of any of the following: lipid-lowering drugs or total cholesterol >240 mg/dl, triglycerides (TG) >150 mg/dl, low-density lipoprotein >130 mg/dl, and high-density lipoproteins (HDL) <50 mg/dl for female and <40 mg/dl for male. Diabetes Mellitus was defined as symptoms of diabetes, fasting blood sugar >126 mg/ dl or HbA1C level > 6.5 or if a patient was on oral hypoglycemic agents. Hypertension was defined as systolic blood pressure >140 mmHg and/or diastolic blood presssure >90 mmHg and/or use of anti-hypertensive treatment.[6,7]

Significant CAD was defined as a diameter stenosis >50% in each major epicardial artery. Normal vessels were defined as the complete absence of any disease in the left main coronary artery (LMCA), left anterior descending (LAD), right coronary artery (RCA), and left circumflex (LCX) as well as in their main branches (diagonal, obtuse marginal, ramus intermedius, posterior descending artery, and posterolateral branch). The patients were classified as having single-vessel disease (SVD), double-vessel disease (DVD) or triple vessel disease (TVD).

Data were collected retrospectively from January 1, 2022 to December 30, 2022 regarding demographic variables, clinical presentation, duration of symptoms, risk factors (age, smoking, hypertension, diabetes mellitus, alcohol consumption, dyslipidemia, and family history), left ventricular ejection fraction, renal function test, cardiac biomarker, angiographic profile, complication of angiography, and in-hospital outcomes were retrieved from the patients’ case note from the medical department of the hospital and electronic data registry system (Midas) in the proforma.

The data obtained were entered into Microsoft Excel 2013 and analyzed using IBM SPSS Statistics version 21. Point estimate and 95% confidence interval were calculated.

## Results

Among 192 patients, 129(67.18%) were male with male to female ratio of 2.04. The mean age of the patients was 61.29 ± 12.50 years with most patients 59 (30.72%) lying within the range of 61-70 years “Table 1”.

**Table 1.**
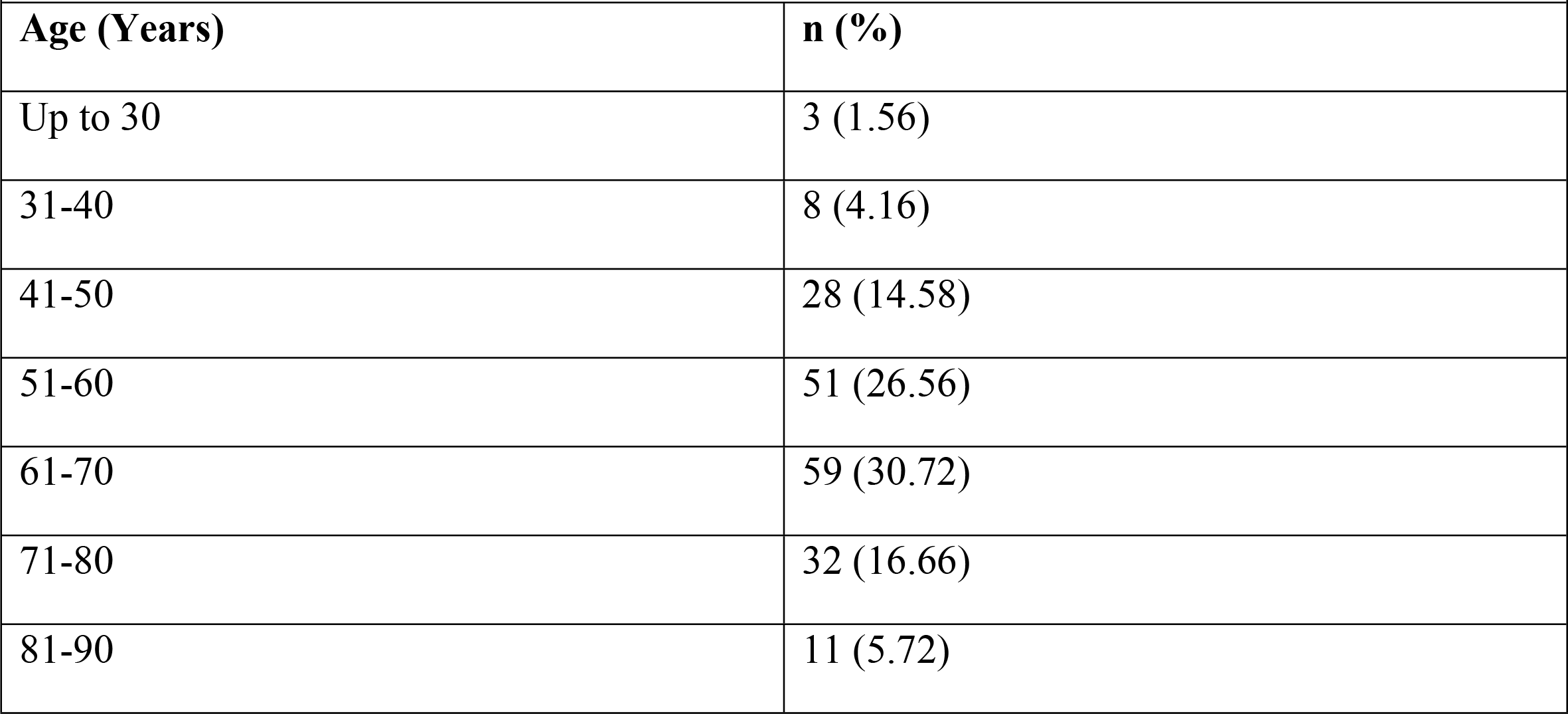
Age-wise distribution of patients with ACS (n= 192)

| **Table 1. Age-wise distribution of patients with ACS (n= 192)** |  |
| --- | --- |
| **Age (Years)** | **n (%)** |
| Up to 30 | 3 (1.56) |
| 31-40 | 8 (4.16) |
| 41-50 | 28 (14.58) |
| 51-60 | 51 (26.56) |
| 61-70 | 59 (30.72) |
| 71-80 | 32 (16.66) |
| 81-90 | 11 (5.72) |

Among total Of 192 patients, 77 (40.10%) had triple vessel disease (TVD) and the most commonly involved vessel was left anterior descending artery (LAD) which was present in 157 (81.77%) patients “Table 2”.

**Table 2.** Distribution of patients according to angiographic finding (n= 192)

|  |  |
| --- | --- |
| **Number of vessels involved** | **n (%)** |
| SVD | 51 (26.56) |
| DVD | 64 (33.33) |
| TDV | 77 (40.10) |
| **Type of vessel involved** |  |
| LAD | 157 (81.77) |
| RCA | 144 (75) |
| LCX | 101 (52.60) |
| OM | 8 (4.16) |
| LM | 4 (2.08) |

Electrocardiography (ECG) and cardiac biomarkers were used to determine the type of ACS. ECG showed that ST-elevation myocardial infarction (STEMI) was the most common type of ACS which was present in 134 (69.79%) of the total patients “Fig 1”. Cardiac troponin I (cTnI) is a cardiac biomarker for ischemic heart disease. The minimum and maximum values of cTnI was 0.1 and 303.11 respectively with a mean value of 12.89±33.53. Similarly, echocardiography was performed to determine the left ventricular ejection fraction (LVEF) which was used to determine the systolic function of the heart where the minimum and maximum recorded values were 10% and 62% respectively with a mean of 44.19±12.66%. Single-vessel disease is common in unstable angina however, triple-vessel disease is more common in patients with STEMI and NSTEMI “Fig 1”

**Fig 1:**
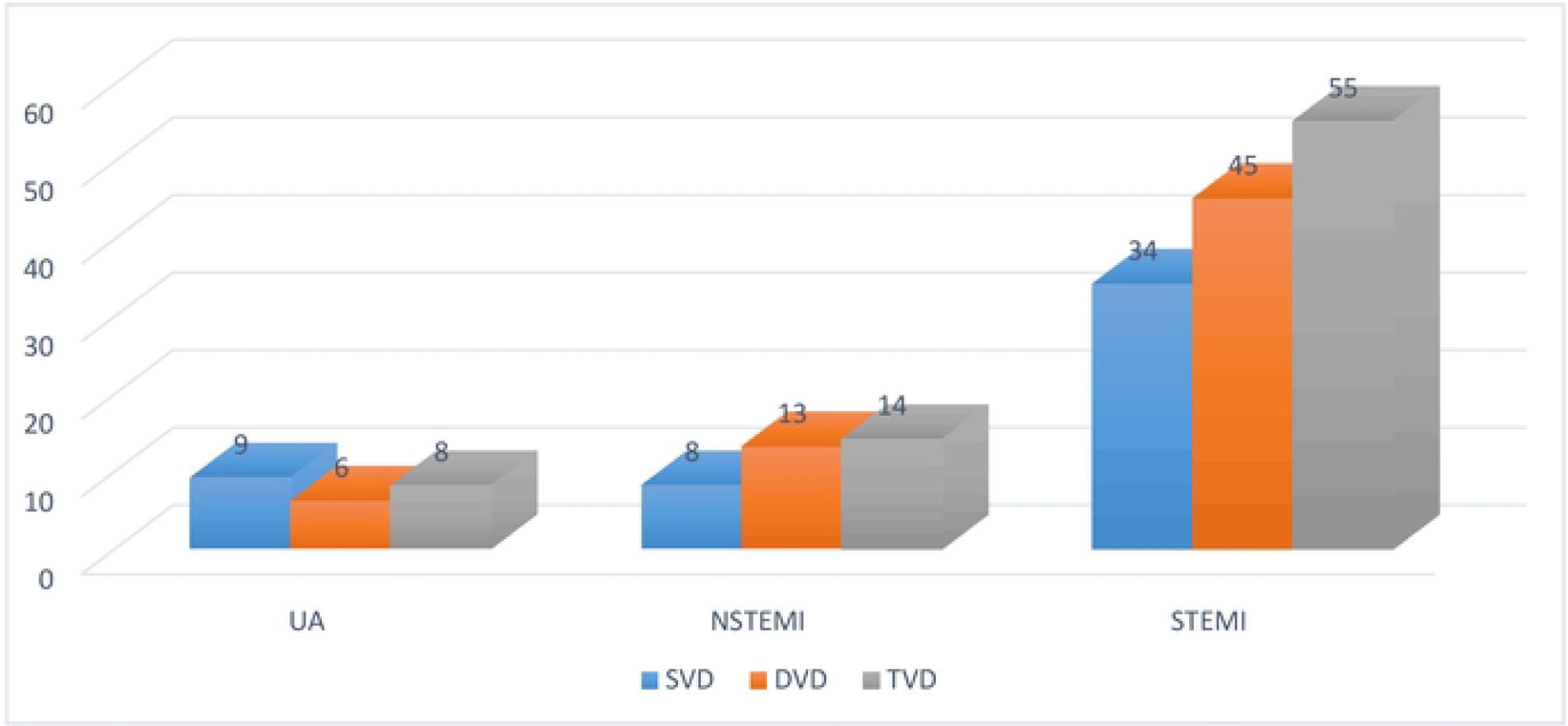
Distribution of coronary vessel involvement in patients with ACS (n=l92)

In our study, there were 16 (8.33%) young patients (< 45 years). The incidence of double vessel disease was higher in younger patients whereas triple vessel disease was more common in elderly patients (>45 years) “Fig 2”

**Fig 2:**
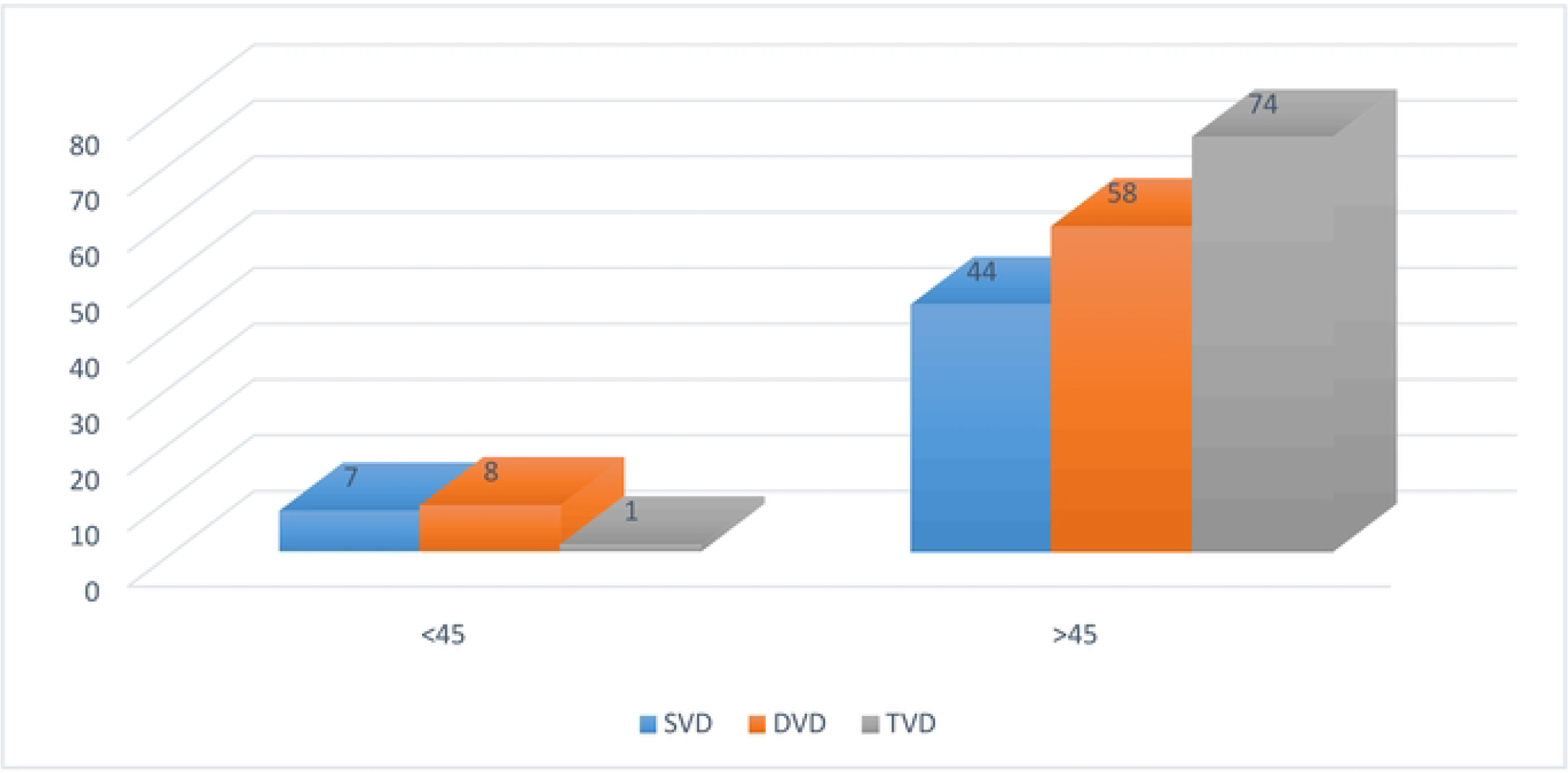
Distribution of coronary vessel according to age (n=l92)

Chest pain was the most common presentation which was present in 163 (84.89%) patients followed by shortness of breath 15 (7.81%) “Table 3”

**Table 3.**
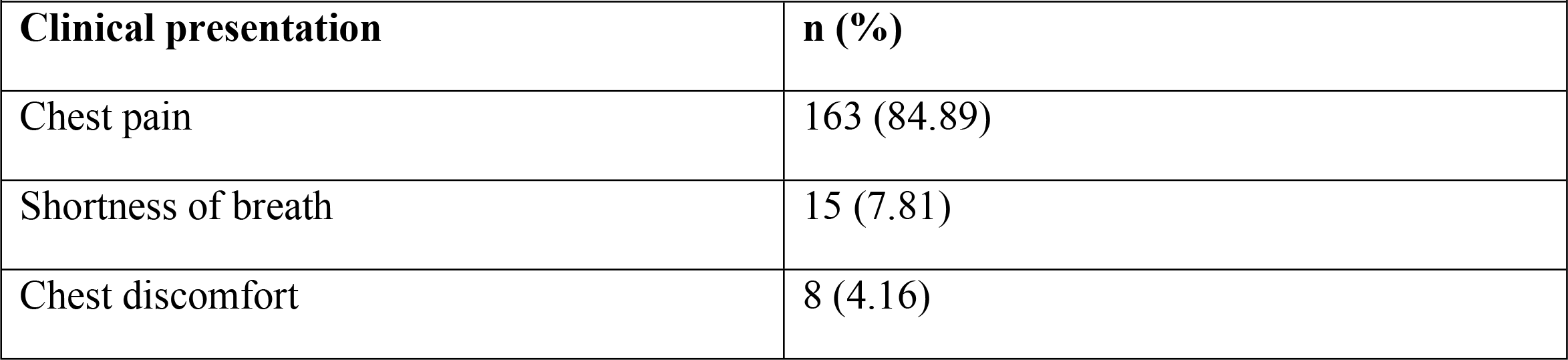
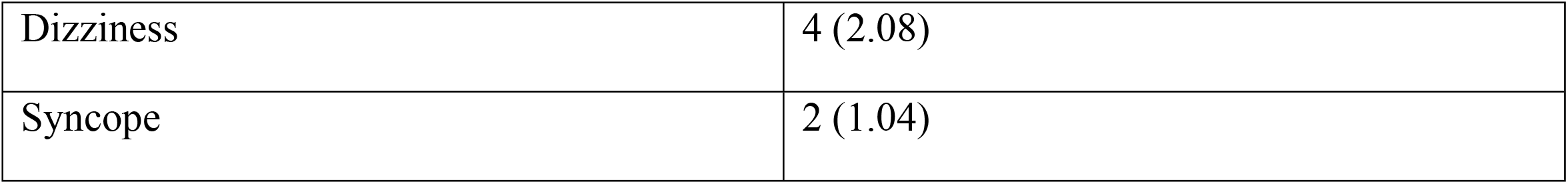
Distribution of patients according to clinical presentation (n= 192)

|  |  |
| --- | --- |
| **Clinical presentation** | **n (%)** |
| Chest pain | 163 (84.89) |
| Shortness of breath | 15 (7.81) |
| Chest discomfort | 8 (4.16) |
| Dizziness | 4 (2.08) |
| Syncope | 2 (1.04) |

Among 192 patients, diabetes mellitus and hypertension were present in 122 (63.54%) and 97 (50.52%) patients respectively followed by dyslipidemia 68 (35.41%) “Table 4”

**Table 4.** Distribution of patients according to underlying condition (n= 192)

|  |  |
| --- | --- |
| **Underlying condition** | **n (%)** |
| Diabetes mellitus | 122 (63.54) |
| Hypertension | 97 (50.52) |
| Dyslipidemia | 68 (35.41) |
| Smoking | 57 (29.68) |
| Alcohol | 32 (16.66) |
| Family history | 4 (2.08) |

In our study, inferior wall was the most commonly involved 77 (40.10%) followed by anterior wall 47 (24.47%) and anteroseptal wall 33 (17.18%) respectively “Fig 3”

**Fig 3:**
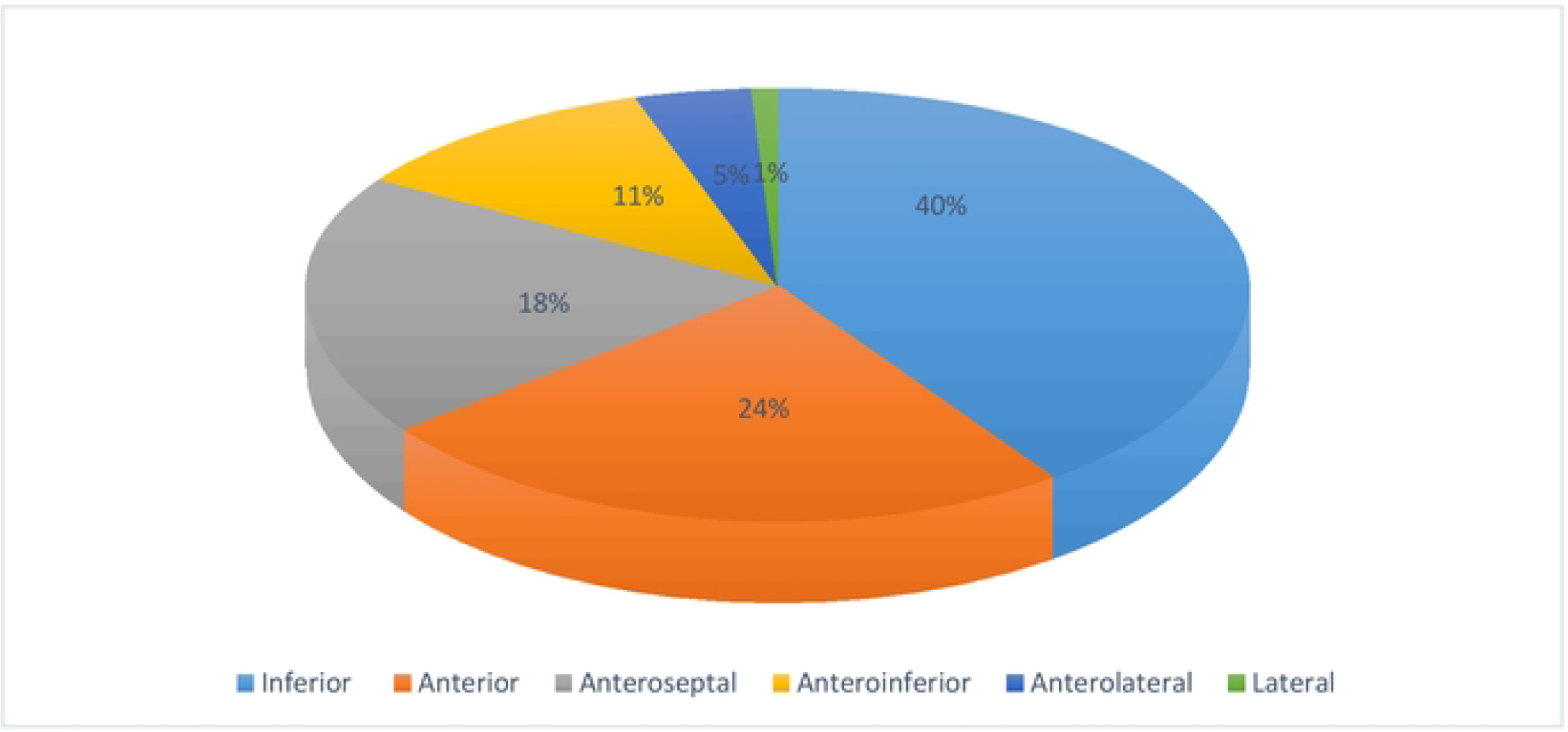
Distribution of study population according to the wall involvement (n=192)

Out of 192 patients, 44 (22.91%) patients developed complications. Among 44 patients, arrhythmia 18 (40.90%) was the most common complication followed by heart failure 12 (27.27%) “Fig 4”. The duration of hospital stays ranged from 1 to 13 days with a mean duration of 4.42±1.87 days. Ten patients died during hospital stay accounting an incidence of 5.20%.

**Fig 4:**
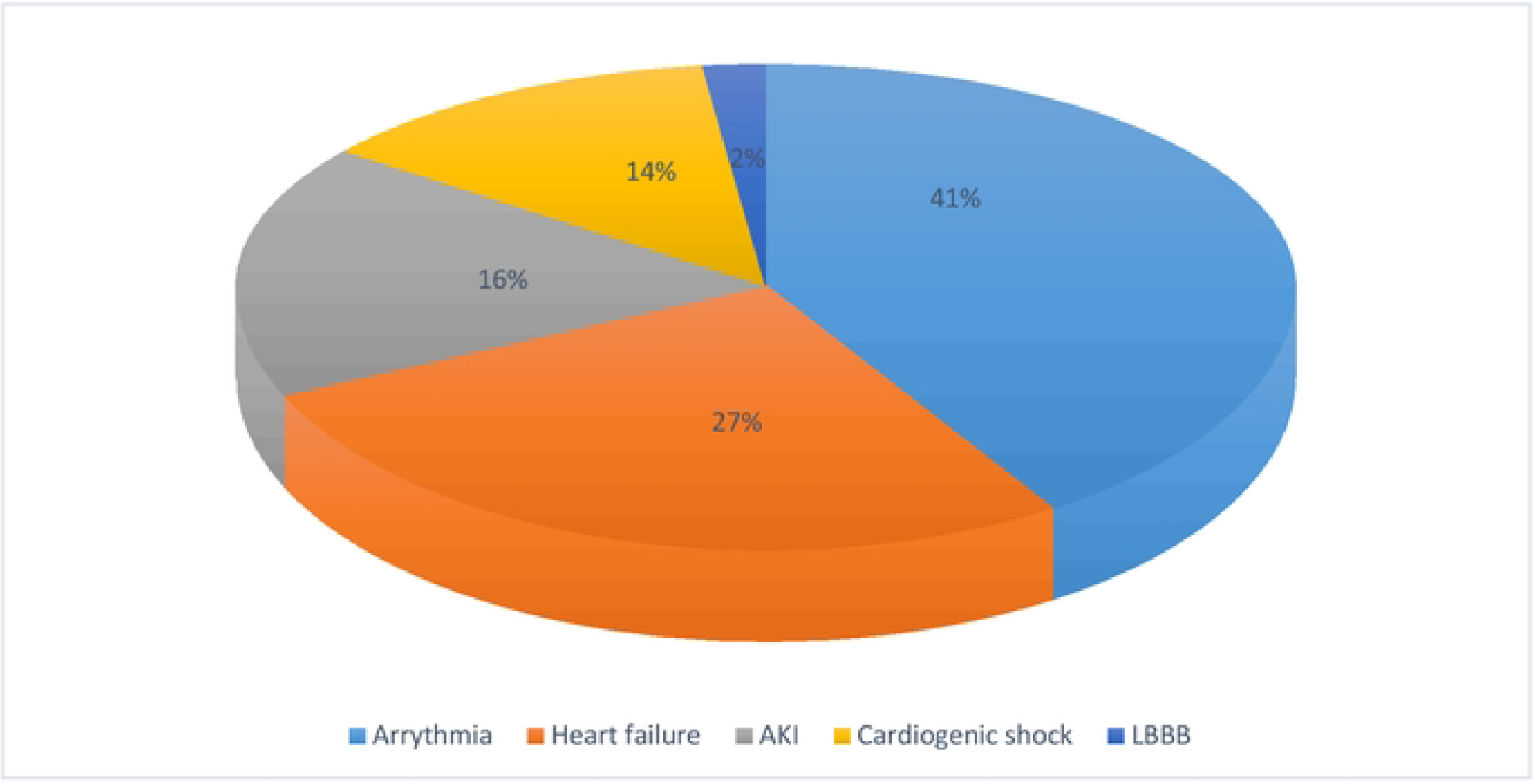
Distribution of study population according to complications (n=44)

## Discussion

Ischemic heart disease followed by stroke is the leading cause of death worldwide of which 80% of deaths and 85% of disabilities occur from cardiovascular disease in developing countries.[8-10]

In our study, the mean age of presentation was 61.29 years which was comparable to studies done in Nepal, India, and USA.[11-16] The incidence of ACS in young (<45 years) ranges from 5-10%.[17-19] Among 192 patients, young patients (<45 years) comprises 16 (8.33%) which was similar to another study done in Nepal.[20]

Angiographic findings in our study revealed more involvement of triple vessel 77 (40.10%) than double vessel disease 64 (33.3%) which was comparable to studies done in Pakistan and Bangladesh.[21,22] However, other studies done had a high prevalence of single vessel disease.[23-26] The high incidence of triple vessel disease, our study might be due to high frequency of diabetes mellitus as a risk factor. In this study, TVD was more common in patients with STEMI and NSTEMI whereas SVD was more common in patients with UA. The left anterior descending artery (LAD) was most the commonly involved followed by the right coronary artery (RCA) which was comparable to a study conducted in India[25]

Echocardiographic finding showed that STEMI 134 (69.79%) was most common presentation among the patients with ACS followed by NSTEMI 35 (18.22%), which was concordant with the study conducted in Nepal[26] and India[25]. However, studies conducted in European countries typically present NSTEMI[27-29]. ECG finding revealed inferior wall MI 77 (40.10%) was most common followed by anterior wall 47 (24.47%) which was comparable, with the study conducted in Pakistan.[30]

Diabetes mellitus, smoking, hypertension, and dyslipidemia are established risk factors for coronary artery disease. In our study, diabetes mellitus was the most common risk factor which was prevalent in 122 (63.54%) of total patients which was much higher than other studies conducted in Nepal[26,31], INTERHEART, GRACE, ACTION, EHS, and PACIFIC registries[32-26]. Diabetes mellitus was followed by hypertension which was present in 97 (50.52%) which was comparable to studies done in Nepal[26], India[37], Pakistan[38], and GRACE[33] registry but lower than ACTION[34] and PACIFIC[36]registry.

In our study, among 192 patients, 44 (22.91%) patients developed complications. Among them, arrhythmia 18 (40.90%) was most common followed by heart failure 8 (18.18%). This complication rate was similar to that reported in a study conducted in Nepal and India[39].

In this study, 10 (5.20%) patients died during their hospital stay and all were due to STEMI which was similar to the GRACE registry.

This was a single-center, descriptive cross-sectional study within a limited time frame. We could not calculate the magnitude of the disease and correlate the different risk factors with angiographic findings. A nationwide multicenter prospective study should be conducted to determine the magnitude of the disease.

## Conclusion

In our study, triple vessel disease was the most common angiographic finding among patients with acute coronary syndrome. Diabetes mellitus was the most common modifiable risk factor which could be the reason for multiple vessel involvement. Arrhythmia 44 (40.90%) was the most common complication developed. 10 (5.20 %) patient died during the hospital stay. Health policy makers must emphasize primary preventive measures, while clinicians/physicians should focus on primary and secondary preventive measures to decrease morbidity and mortality related to coronary artery disease.

## Data Availability

Data will be provided if necessary upon request by the editors.

## Conflict of interest

**None**

## Notes

### Competing Interest Statement

The authors have declared no competing interest.

### Funding Statement

Unfunded study

### Author Declarations

Approval for study was done by Institutional Review Committee (IRC) of Chitwan Medical College with a reference number of 079/080-161.

